# Systemic Metabolic Alterations after Aneurysmal Subarachnoid Hemorrhage: A Plasma Metabolomics Approach

**DOI:** 10.1101/2025.01.06.25320083

**Authors:** Bosco Seong Kyu Yang, Jude P.J. Savarraj, Hua Chen, Sarah N. Hinds, Glenda L. Torres, Alice S. Ryan, Folefac D. Atem, Philip L. Lorenzi, Xuefang S. Ren, Neeraj Badjatia, Huimahn A. Choi, Aaron M. Gusdon

## Abstract

**Background:** Aneurysmal subarachnoid hemorrhage (aSAH) causes systemic changes that contribute to delayed cerebral ischemia (DCI) and morbidity. Circulating metabolites reflecting underlying pathophysiological mechanisms warrant investigation as biomarker candidates.

**Methods:** Blood samples, prospectively collected within 24 hours (T1) of admission and 7-days (T2) post ictus, from patients with acute aSAH from two tertiary care centers were retrospectively analyzed. Samples from healthy subjects and patients with non-neurologic critical illness served as controls. A validated external analysis platform was used to perform untargeted metabolomics. Bioinformatics analyses were conducted to identify metabolomic profiles defining each group and delineate metabolic pathways altered in each group. Machine learning (ML) models were developed incorporating key metabolites to improve DCI prediction.

**Results:** Among 70 aSAH, 30 healthy control, and 17 sick control subjects, a total of 1,117 metabolites were detected. Groups were matched among key clinical variables. DCI occurred in 36% of aSAH subjects, and poor functional outcome was observed in 70% at discharge. Metabolomic profiles readily discriminated the groups. aSAH subjects demonstrated a robust mobilization of lipid metabolites, with increased levels of free fatty acids (FFAs), mono- and diacylglycerols (MAG, DAG) compared with both control groups. aSAH subjects also had decreased circulating amino acid derived metabolites, consistent with increased catabolism. DCI was associated with increased sphingolipids (sphingosine and sphinganine) and decreased acylcarnitines and S-adenosylhomocysteine at T1. Decreased lysophospholipids and acylcarnitines were associated with poor outcomes. Incorporating metabolites into ML models improved prediction of DCI compared with clinical variables alone.

**Conclusions:** Profound metabolic shifts occur after aSAH with characteristic increases in lipid and decreases in amino acid metabolites. Key lipid metabolites associated with outcomes (sphingolipids, lysophospholipids, and acylcarnitines) provide insight into systemic changes driving secondary complications. These metabolites may also prove to be useful biomarkers to improve prognostication and personalize aSAH care.

## Introduction

Subarachnoid hemorrhage occurring due to the rupture of an intracranial aneurysm (aSAH) affects approximately 50,000 people per year in the United States^1^. Although the incidence of aSAH is considerably lower than other forms of stroke, those affected are typically younger (average 40-60 years of age), with the resultant high morbidity and mortality leading to a disproportionate number of productive years of life lost^2^.

In addition to early brain injury occurring within the first few days after aneurysm rupture, 30% of aSAH patients develop delayed cerebral ischemia (DCI)^3^. DCI often results in new neurological deficits and contributes to worse clinical outcome^4^. Although early detection of delayed complications such as DCI is of critical importance, prediction remains difficult, with clinical and radiographic scores being of limited use^5,6^. There is a lack of validated biomarkers able to predict delayed complications and outcomes after aSAH^7^.

Metabolites may provide key insights into the pathophysiological changes contributing to DCI after aSAH. We have previously demonstrated that tricarboxylic acid (TCA) cycle metabolites are associated with systemic inflammation and outcomes after aSAH^8^. However, lipid metabolites likely play key roles in the pathophysiology leading to DCI and poor functional outcomes after aSAH. Lipid metabolites such as sphingosine-1-phosphate (S1P) and lysophosphatidic acid (LPA) are vasoactive and may contribute to the development of vasospasm and DCI^9–11^. The only genome-wide association study (GWAS) considering outcomes after aSAH found a significant association with a variant in *SPNS2* (rs12949158), an S1P transporter that can potentially influence the local concentrations of S1P in the central nervous system^12^.

We hypothesized that circulating metabolites, especially lipids, would provide insight into pathophysiological changes occurring after aSAH and be able to predict DCI and functional outcomes. We performed untargeted metabolomics using plasma samples from patients with aSAH as well as controls in order to identify metabolites that may serve as clinically relevant biomarkers.

## Methods

### Subjects

Patients were included from two sites: 1) the University of Texas Health Science Center (UTHSC), Memorial Hermann Hospital (MHH)-Texas Medical Center, and 2) the University of Maryland School of Medicine (UMSOM). Patients admitted to the neurocritical care unit at MHH were prospectively enrolled in a biobank between July 2019 and August 2022. Patients admitted at UMSOM were selected from those enrolled in the INSPIRE phase 2 randomized controlled trial (NCT03201094) who had available plasma samples^13^. Detailed methods are available in the published trial results^13^. Inclusion criteria included: age 18 years or older, spontaneous aSAH admitted within 24h of ictus, and confirmed aneurysmal source on either computed tomography angiogram (CTA) or digital subtraction angiogram (DSA). Exclusion criteria included subarachnoid hemorrhage from a non-aneurysmal etiology (e.g. trauma, arteriovenous malformations, or mycotic aneurysms) and comorbidities affecting baseline inflammation and circulating metabolites (e.g. autoimmune disorders and active malignancy). Two types of control subjects were included: 1) healthy controls recruited at the University of Texas Physician Cardiology Clinic during their routine follow-up and 2) critically ill patients admitted to the ICU at MHH but without neurological injury.

### Ethical approval

Institutional review boards (IRB) at both institutions reviewed and approved this study. IRB approvals were as follows: UTHSC-MHH (HSC-MS-12-0637) and UMSOM (HP-00074174). All methods were performed in accordance with the Declaration of Helsinki. Prior to enrollment, informed consent was obtained from all patients or their legal guardians.

### Clinical data

For each subject, data were prospectively collected including demographic and clinical data including age, sex, ethnicity, and clinical severity determined by the Hunt-Hess Scale (HHS) score. DCI was defined as clinical deterioration [new focal neurological deficit or decrease of at least two points on the Glasgow Coma Scale (GCS)] not otherwise explained or a new infarct seen on neurological imaging^14^. Adjudication for DCI required the consensus of at least two neurointensivists. Functional outcomes were assessed using the modified Rankin Scale (mRS) at discharge as assessed by the attending on service with trained research personnel performing follow-up phone calls at 3-months and administering standardized questionnaires. Good functional outcomes were defined as mRS 0-3 and poor functional outcomes were defined as 4-6.

### Biosamples

Blood samples were collected from subjects in ethylenediaminetetraacetic acid (EDTA) containing tubes. Samples were centrifuged at 4°C and stored at -80°C until analysis. Two time points were used for metabolomics analyses: within 24-hours of admission while subjects remained nil per oral for interventional procedures (T1) and at post-bleed day 7 (T2).

### Metabolomics

Plasma samples (200 μL) were sent to Metabolon (Morrisville, NC, USA) for untargeted metabolomics analysis in a single batch. Detailed descriptions of the metabolomics platform, which consists of four independent ultra-high-performance liquid chromatography-tandem mass spectrometry (UPLC-MS/MS) instruments and methods have been published elsewhere^15–17^. Median and standard deviation of internal standards were used to assess instrument variability. Identification of each metabolite was accomplished by automated comparison of each ion to a standard library. Areas under the curve (AUC) were calculated for each peak. Raw AUC values were normalized by correcting for between-day variation in instrument calibration using internal standards and median values for each run day. Missing values were imputed using k-nearest neighbors with 10 neighbors used for each imputation. All results were subsequently logarithmically transformed.

### Bioinformatics

Fold changes (FC) were calculated for each metabolite considering different comparisons: 1) aSAH vs healthy controls, 2) aSAH vs. sick controls, 3) aSAH with vs. without DCI, and 4) aSAH with good vs poor functional outcomes at discharge. Changes in metabolites were considered to be significantly increased if FC > 2 and decreased if FC < 0.5 with false discovered rate (FDR) corrected *p*-values < 0.05. FCs and corresponding *p-*values were calculated using the R package MetaboAnalyst 5.0 (https://www.metaboanalyst.ca)^18^ Next, sparse partial least squared discriminant analysis (sPLS-DA, a supervised learning technique) from the mixOmics library in R (http://mixomics.org) was used to identify metabolites that distinguish among different groups. Variable importance in Projection (VIP) scores were calculated for the metabolites to visualize their relative importance in group discrimination. Gene ontology (GO) and Kyoto Encyclopedia of Gene and Genomes (KEGG) enrichment analysis were used to analyze the differences in biological pathways^19^. Agglomerative hierarchical clustering, a form of unsupervised learning, using the Euclidean distance as a similarity measure and Ward’s linkage as a clustering algorithm was performed to identify patterns of metabolites distinguishing groups ^22^.

### Machine learning (ML) models

ML-based prediction models can learn from high-dimensional data to find hidden features that improve predictions^20^. We tested the additive predictive value of the identified metabolites for DCI and poor functional outcomes at discharge. Two sets of variables were used. The baseline variables contained age and HHS, and the extended set included the metabolites significant for each outcome in addition to the baseline variables. The models based on elastic net logistic regression with equal L1 and L2 regularization (ELR) ^21^ and Extreme Gradient Boosting models (XGB)^22^ were utilized. The two models were chosen to employ regularization to limit overfitting and examine both linear and non-linear associations between the predictors and outcomes. To reliably assess each model’s performance, five-fold cross-validation was used, and the area under the receiver operating characteristic (AUROC) curve for all folds were combined to derive the 95% confidence interval (CI) for the AUROC for each model. For XGB models, feature importances were analyzed with mean decrease in impurity (MDI) and Shapley additive explanation (SHAP) values^23^. While the former allowed relative quantification of each feature’s global impact on the trained model, the latter allowed analysis of more granular aspects of each feature’s influence on the prediction. ML models were developed and evaluated using Python (v3.11) using the ‘sci-kitlearn’ toolbox.

### Statistics

Patient characteristics were compared using the Wilcoxon rank sum test for continuous variables and Chi-square test and Fisher’s exact test for categorical variables as necessary. For the statistical tests, two-tailed p-values were calculated, and a p-value of 0.05 was used as the threshold for statistical significance. Bioinformatics and statistical analyses were performed with R 4.3.1 (R Core Team, 2023).

## Results

### Patient population

Among 117 patients finally included in the study, 46 aSAH patients, 15 healthy controls, and 17 hospitalized controls were recruited from UTHSC, and 24 aSAH patients and 15 healthy controls were recruited from UMSOM. The median ages for aSAH patients, hospitalized controls, and healthy controls were 56, 58, and 57, respectively (p = 0.44); 59%, 41%, and 50% were females, respectively (p = 0.31). Among aSAH patients, the median Hunt-Hess scale (HHS) was 3 [Inter Quartile Range (IQR): 3-4]. 36% DCI occurred in 36% of patients, and poor outcomes were observed in 70% at discharge but only 24% by 3-months after aSAH (Table 1). 8 (17%) aSAH patients recruited from UTHSC had a history of diabetes mellitus. For the hospitalized controls, the major reasons for their admission to the intensive care unit were respiratory failure due to acute exacerbation of chronic obstructive lung diseases (COPD) (35%) and pneumonia (24%). Other reasons included acute decompensated heart failure and septic shock. In comparison, none of aSAH patients recruited from UTHSC had a history of COPD.

**Table 1.** Baseline characteristics.

|  | Healthy Controls | Hospitalized Controls | aSAH | *P*-Value* |
| --- | --- | --- | --- | --- |
| N | 30 | 17 | 70 |  |
| Age [median (IQR)] | 57 (52-70) | 58 (41-70) | 56 (47-65) | 0.44 |
| Sex [Female (%)] | 15 (50%) | 7 (41%) | 42 (59%) | 0.31 |
| Race [N (%)] |  |  |  |  |
| Black | 9 (30%) | 7 (41%) | 18 (26%) | 0.64 |
| White | 20 (67%) | 9 (53%) | 49 (70%) |  |
| Asian | 1 (3%) | 1 (6%) | 3 (4%) |  |
| Ethnicity [N (%)] |  |  |  |  |
| Hispanic | 6 (20%) | 4 (24%) | 27 (39%) | 0.14 |
| HHS [median (IQR)] |  |  | 3 (3-4) |  |
| DCI [N (%)] |  |  | 25 (36%) |  |
| mRS discharge 0-3 [N (%)] |  |  | 49 (70%) |  |
| mRS 3-months 0-3 [N (%)] |  |  | 17 (24%) |  |

\**P*-Values reflect comparisons between the three groups. Abbreviations: IQR (Interquartile range), HHS (Hunt Hess Scale), mRS (modified Rankin Scale)

### Metabolites

A total of 1,383 metabolites were detected from plasma samples. 268 metabolites were either unidentified metabolites or medications and were removed from further analysis, resulting in 1117 metabolites. Metabolites consisted of 485 lipids, 215 amino acids, 39 cofactors and vitamins, 35 nucleotides, 33 peptides, 23 carbohydrates, 15 energy-related pathways (glycolysis, gluconeogenesis, and the TCA cycle), as well as partially characterized molecules.

### Supervised approach in finding discriminative metabolites

Based on the samples collected at T1, sPLS-DA was utilized to identify metabolites distinguishing aSAH from sick or healthy controls. Five-fold cross-validation was used, resulting in an R2Y value of 0.96, Q2 values of 0.89, and an overall accuracy of 0.99. The scores plot demonstrates that the top 15 metabolites readily distinguished aSAH subjects from healthy controls, while there was more overlap between sick controls and aSAH subjects (Figure 1A). The top 15 metabolites distinguishing the three groups as well as their direction of change are shown in the corresponding loading plot (Figure 1B). Enrichment analysis was conducted to identify pathways that are altered comparing aSAH and healthy control subjects (Figure 1C) or aSAH and sick control subjects (Figure 1D). aSAH subjects demonstrated increases in catabolic pathways involving protein breakdown and amino acid metabolism as well as fatty acid biosynthesis. Changes in individual metabolites are shown in volcano plots comparing FC for aSAH vs healthy controls (Figure 1E) and aSAH vs sick controls (Figure 1F). A total of 90 metabolites were significantly increased, while 63 were significantly decreased comparing aSAH and healthy controls (Figure 1E, Table S1). A total of 68 metabolites were significantly increased, while 71 were significantly decreased comparing aSAH and sick controls (Figure 1F, Table S2). aSAH subjects demonstrated notable increases in several classes of lipid metabolites, while amino acid metabolites were prominently decreased compared with controls. TCA cycle metabolites were also decreased in aSAH compared with controls.

**Figure 1:**
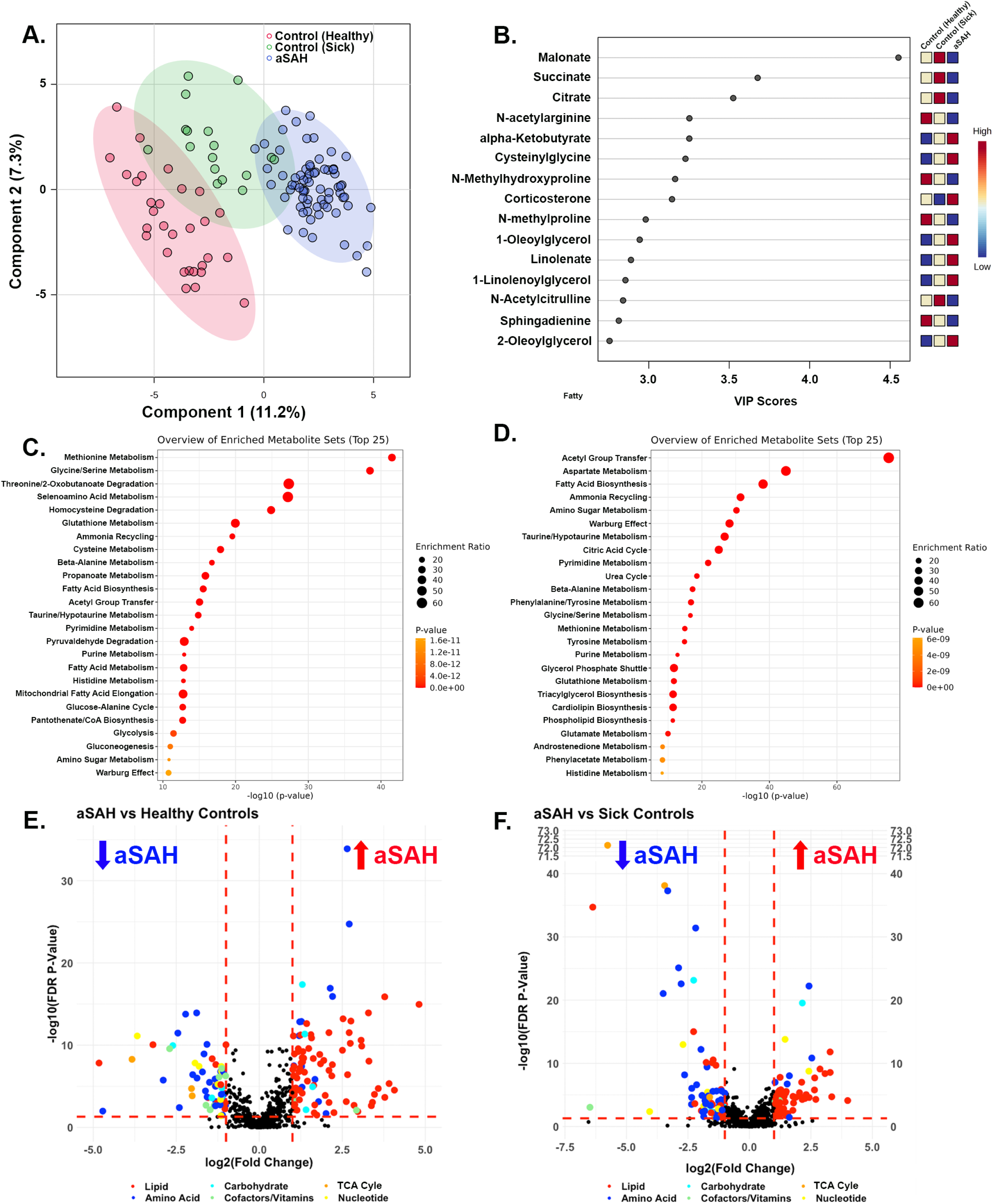
Changes in metabolites comparing controls and aSAH. Metabolomic profiles effectively discriminate aSAH, sick control, and healthy subjects using sparse partial least squared discriminant analysis (sPLS-DA) (A). VIP scores plot identifies the 15 most important metabolites in the classification (B). Enrichment analysis results showing significantly altered metabolic pathways in aSAH subjects compared with (C) healthy control (C) and sick control (D) subjects. Volcano plots are shown demonstrating changes in each individual comparing aSAH with healthy controls (E) and sick controls (F). Metabolites are color-coded according to category. Vertical dashed lines denote fold changes of 2 and 0.5 (log_2_ transformed 1 and -1). The horizontal dashed line denotes an FDR-corrected p-value of 0.05.

### An unsupervised approach to confirm the discriminative metabolites

Given the differences in lipid metabolites noted in Figure 1, agglomerative HC was used to confirm the ability of lipid metabolites to distinguish aSAH from healthy and sick controls. aSAH patients were readily differentiated from healthy controls (Figure 2A) and sick controls (Figure 2B) based on levels of lipid metabolites. In particular, free fatty acids (FFAs) (long-chain, poly-, and mono-unsaturated) as well as mono- and diacylglycerol were higher in aSAH patients compared to healthy and sick controls. Functional analysis of the fatty acids and acyl glycerols with significant changes in their expression levels revealed they were involved in multiple pathways including regulation of lipolysis, ferroptosis, bile secretion, PI3K-Akt signaling pathway, and steroid hormone biosynthesis (Figure S1).

**Figure 2:**
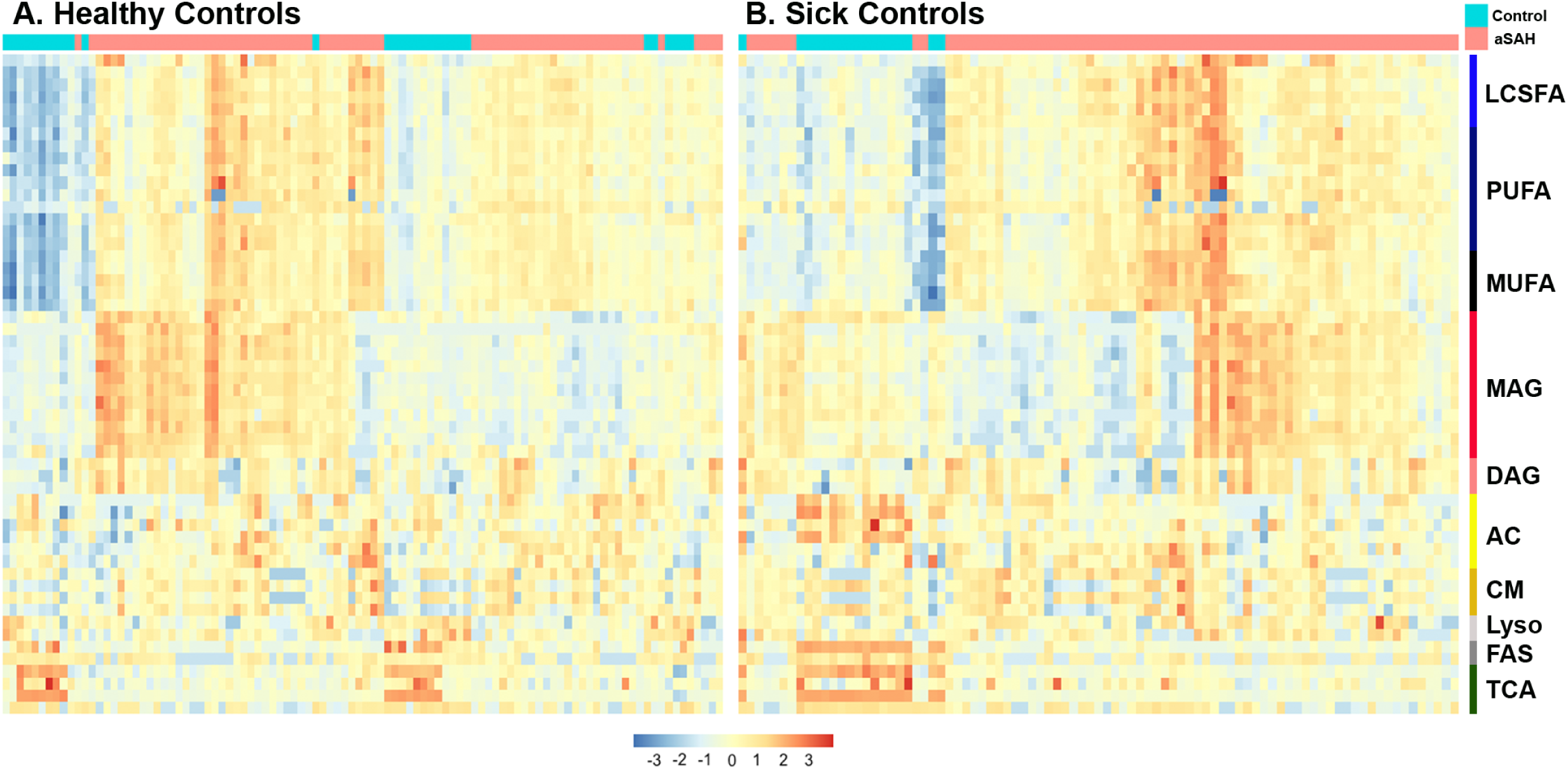
Heatmaps demonstrating changes in lipid metabolites comparing aSAH and controls. Unsupervised clustering of aSAH vs healthy control (A) and sick control (B) subjects was performed according to levels of several classes of lipid metabolites to generate heatmaps. Lower relative values are blue while higher relative values are red. Among the lipid metabolites, long-chain fatty acids, poly-, and mono-unsaturated fatty acids, mono- and diacylglycerol were particularly higher in aSAH subjects in comparison to controls. Abbreviations: LCSFA (Long- chain saturated fatty acids), PUFA (poly-unsaturated fatty acids), MUFA (mono-unsaturated fatty acids), MAG (monoacylglycerol), DAG (diacylglycerol), AC (acylcarnitines), CM (ceramides), Lyso (Lysophopholipids), FAS (metabolites involved in fatty acid synthesis), TCA (Tricarboxylic Acid cycle metabolites).

### Metabolites associated with aSAH outcomes

Within aSAH subjects, volcano plots were generated to assess differential levels of metabolites considering DCI (Figure 3A) and discharge mRS (Figure 3B). Subjects with DCI had higher levels of sphingosine and sphinganine and lower levels of S-adenosylhomocysteine, undecanedioate, and octanoylcarnitine compared to subjects without DCI (Figure 3A, Table S3). Subjects with poor functional outcome at discharge had higher levels of N-acetylglutamate, and lower levels of linoleoyl glycerophosphate, linoleoyl glycerophosphocholine, and three acylcarnitines (dihomo-linoleoylcarnitine, eicosenoylcarnitine, and nervonoylcarnitine) compared to subjects with good functional outcome at discharge (Figure 3B, Table S4). These metabolites did not reach significance according to FDR p-value thresholds and were therefore ranked according to raw p-values (Tables S3 and S4).

**Figure 3:**
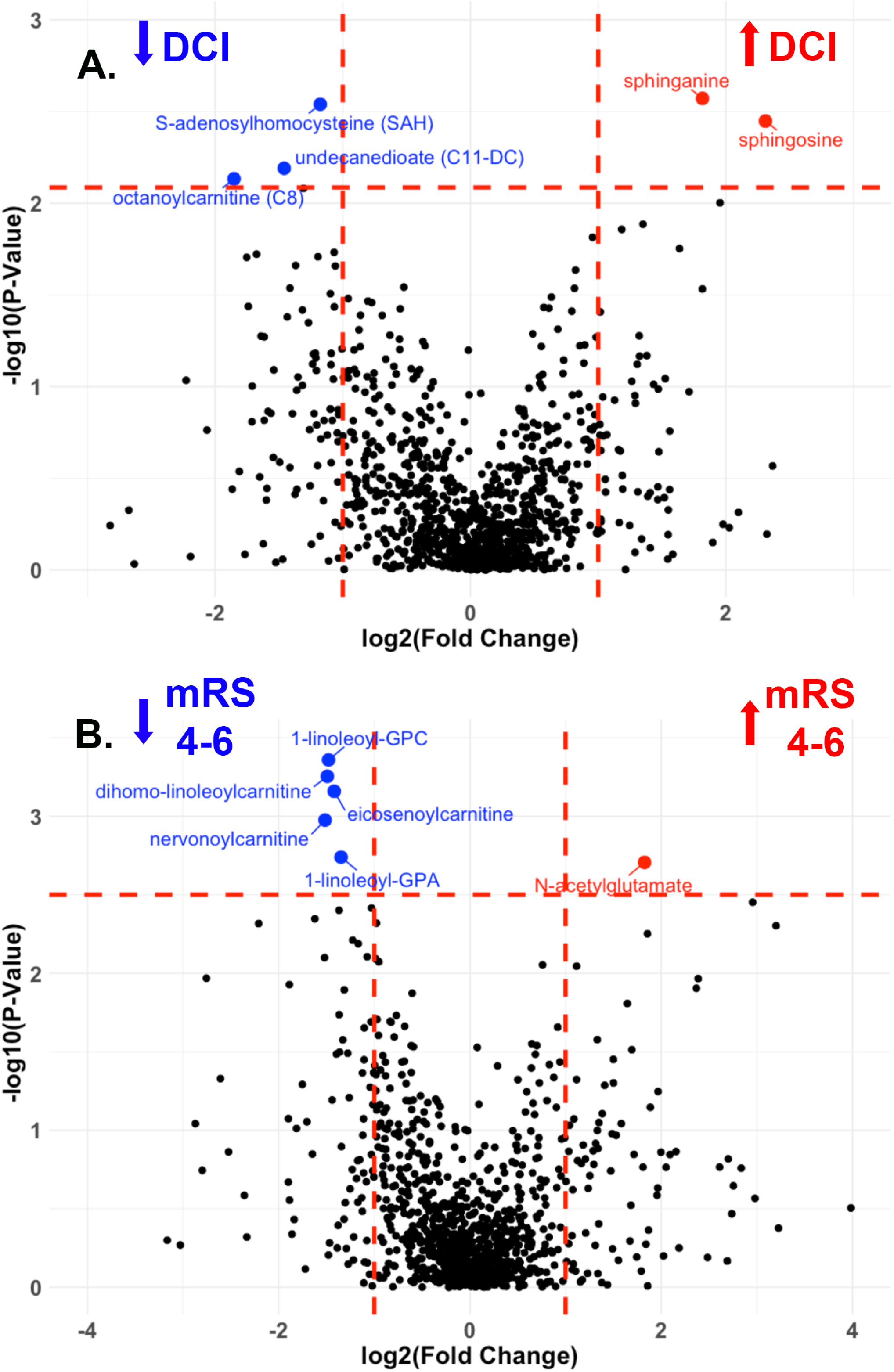
Metabolites showing significant changes in delayed cerebral ischemia and poor functional outcome. Volcano plots demonstrating metabolites with significant changes considering DCI (A) or functional outcome (B). Metabolites are plotted against their raw p-values. Metabolites in red are increased among subjects with DCI (A) and poor outcomes (B), while those in blue are decreased among subjects with DCI and poor outcomes. Vertical dashed lines denote fold changes of 2 and 0.5 (log_2_ transformed 1 and -1). Horizontal dashed lines denote raw p-values of 0.0082 (A) and 0.0032 (B). Poor functional outcomes are defined as mRS 4-5, while good functional outcomes are defined as mRS 0-3 at discharge. Abbreviations: DCI (delayed cerebral ischemia), mRS (modified Rankin Scale) GPC (glycerophosphorylcholine), GPA (glycerophosphate).

Levels of each metabolite found to be associated with outcomes (Figure 3) were depicted as bar plots for all aSAH subjects (N=70), subjects from the UTHSC (N=46), and subjects from the UMSOM (N=24). Considering DCI, levels of each metabolite at T1 were significantly different among subjects with or without DCI when including all subjects (Figure S2A). Considering only subjects from UTHSC, sphingosine (p=0.012) and sphinganine (p=0.023) were significantly higher, while octanoylcarnitine (p= 0.047) was significantly lower among subjects with DCI (Figure S2B). Considering only subjects from UMSOM, octanoylcarnitine (p=0.043) and undecanedioate (p=0.039) were significantly lower among subjects with DCI (Figure S2C). Considering discharge outcomes, among only UTHSC subjects, all six metabolites identified in Figure 3B were significantly different: 1-linoleoyl-GPC (18:2) (p=2.08x10^-4^), 1-linoleoyl-GPA (18:2) (p=0.014), nervonoylarnitine (p=0.011), eicosenoylcarnitine (p=0.003), dihomo- linoleoylcarnitine (p=4.59x10^-4^), and N-acetylglutamate (p=0.005) (Figure S2B). Among UMSOM subjects, nervonoylcarnitine (p=0.019) and dihomo-linoleoylcarnitine (p=0.018) were significantly lower at T1 among subjects with poor outcomes (Figure S2C).

### Changes in the metabolites at early vs late stages of aSAH

Paired comparisons were made considering metabolites early (within 24h of aneurysmal rupture, T1) and late (7 days, T2) after aneurysmal rupture. Changes in individual metabolites are shown in the volcano plot with paired fold changes comparing each metabolite and the late vs early timepoint (Figure 4A). A total of 41 metabolites were significantly increased, while 46 were significantly decreased comparing T2 to T1 (Table S5). Among the metabolites that showed significant changes in levels, 77% were lipids. Most of the others were amino acids. Steroid hormones (androgenic steroids, corticosteroids, pregnenolone steroids, and progestin steroids) decreased from T1 to T2, whereas lipids involved in bile acid metabolism, monoacylglycerols, and diacylglycerols increased. Among metabolites found to be associated with outcomes, subjects with poor outcomes showed significant increases in sphingosine from T1 to T2 (*p*= 0.0056) and a trend toward increased sphingainine (*p*=0.075) (Figure 4B). Although higher acylcarnitines at T1 were associated with better outcomes (Figure 3), subjects with good functional outcomes showed significant decreases in four acylcarnitines from T1 to T2 (Figure 4C): nervonoylcarnitine (*p*=0.039), erucoylcarnitine (*p*=0.0037), docosahaxaenoylcarnitine (*p*=0.0067), adrenoylcarnitine (*p*=0.034).

**Figure 4:**
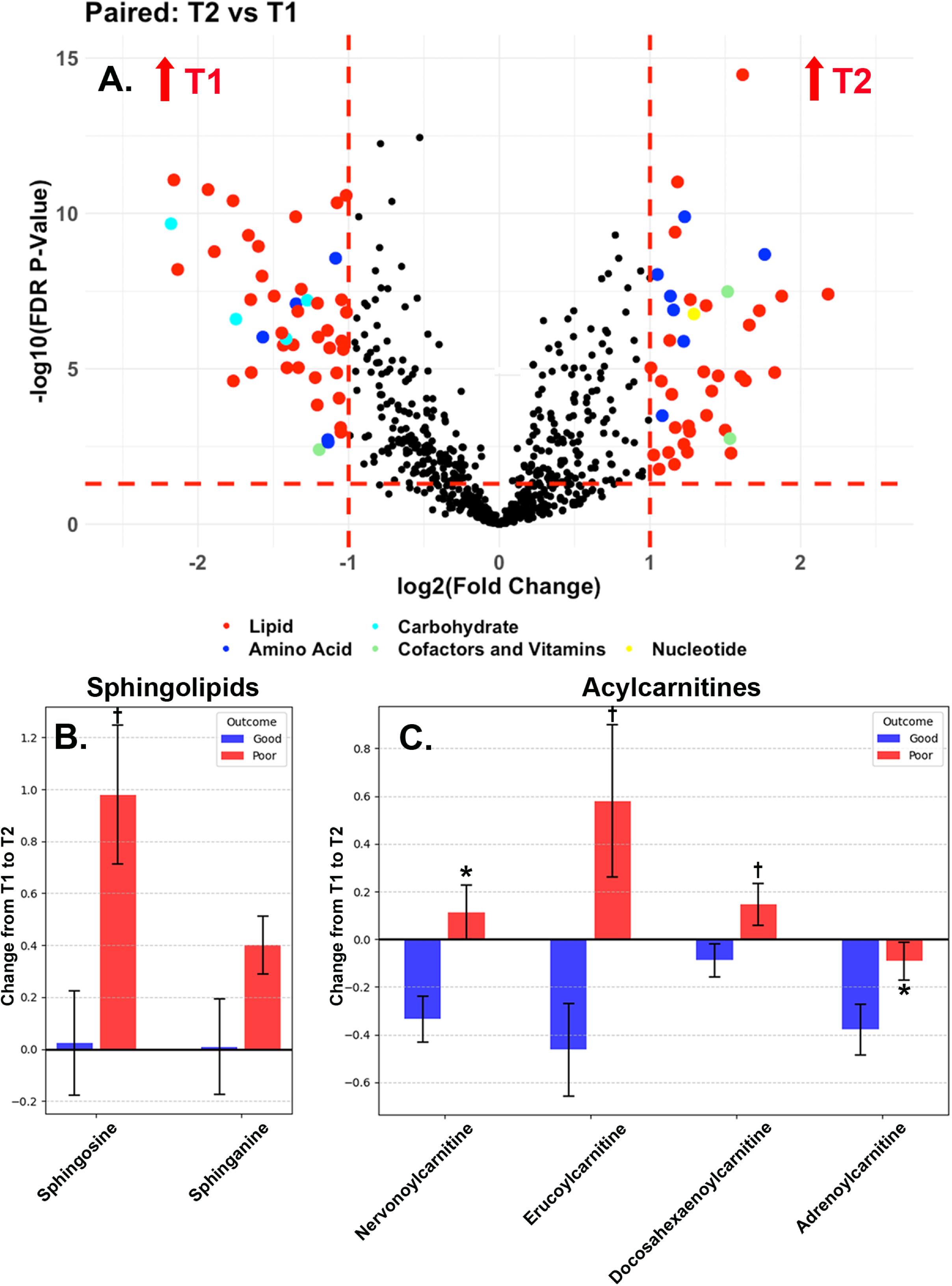
Metabolites showing significant temporal changes after aSAH. Metabolomic profiles were compared between two time points: within 24 hours post ictus (T1) and at 7 days (T2). Volcano plots demonstrate changes in individual metabolites from T1 to T2 (A). Metabolites are color-coded according to category. Vertical dashed lines denote fold changes of 2 and 0.5 (log_2_ transformed 1 and -1). The horizontal dashed line denotes an FDR-corrected p-value of 0.05. Changes in key metabolites from T1 to T2 are shown as bar plots for selected sphingolipids (B) and acylcarnitines (C). Subjects are categorized according to discharge outcome: good (blue, mRS 0-3) and poor (red, mRS 4-6). Error bars represent the standard error of the mean (SEM). Paired t-tests were used to compare subjects with good or poor outcomes for changes in each metabolite with significance denoted as: \**p*<0.05, ^†^*p*<0.01. Abbreviations: mRS (modified Rankin Scale), FDR (false discovery rate).

### Multivariable logistic regression models

Multivariable logistic regression models were developed for metabolites associated with outcomes. Metabolite data were included from T1. After correction for age, clinical severity (HHS), sex, and race, increased sphingosine [OR 9.95 (95%CI 2.42, 59.4)] and sphinganine [OR 4.47 (95%CI 1.54, 20.7)] were associated with the occurrence of DCI (Table 2). Conversely, decreased octanoylcarnitine was associated with the occurrence of DCI [OR 0.20 (95%CI 0.04, 0.65)]. Both age and clinical severity were associated with discharge functional outcome (mRS 0-3: good; mRS 4-6: poor) (Table 2). When controlling for age, clinical severity, sex, and race, decreased 1-linoleoyl-GPC (18:2) [OR 0.17 (95%CI 0.03, 0.76)], dihomo-linoleoylcarnitine [OR 0.20 (95%CI 0.04, 0.77)], and eicosenoylcarnitine [OR 0.19 (95%CI 0.04, 0.76)] were associated with poor functional outcome (Table 2).

**Table 2.** Multivariate Logistic Regression Models for DCI and Discharge Outcome.

| DCI |  |  |  |  |  |  |
| --- | --- | --- | --- | --- | --- | --- |
|  | Sphingosine |  | Sphinganine |  | Octanoylcarnitine |  |
| Variable | OR (CI) | p | OR (CI) | p | OR (CI) | p |
| Metabolite | 9.95<br>(2.42, 59.4) | **0.005** | 4.47<br>(1.54, 20.7) | **0.029** | 0.20<br>(0.04, 0.65) | **0.018** |
| Age | 1.01<br>(0.96, 1.06) | 0.82 | 1.00<br>(0.95, 1.05) | 0.91 | 1.01<br>(0.96, 1.06) | 0.80 |
| HHS | 2.34<br>(0.75, 7.81) | 0.15 | 1.73<br>(0.58, 5.26) | 0.33 | 1.29<br>(0.44, 3.79) | 0.64 |
| Sex | 2.05<br>(0.67, 6.84) | 0.22 | 2.01<br>(0.67, 6.53) | 0.23 | 0.94<br>(0.31, 2.85) | 0.92 |
| Race | 1.09<br>(0.42, 2.92) | 0.86 | 1.11<br>(0.45, 2.87) | 0.82 | 1.28<br>(0.52, 3.28) | 0.59 |
| Discharge Outcome |  |  |  |  |  |  |
|  | 1-linoleoyl-GPC<br>(18:2) |  | Dihomo-linoleoylcarnitine |  | Eicosenoylcarnitine |  |
| Variable | OR (CI) | p | OR (CI) | p | OR (CI) | p |
| Metabolite | 0.17<br>(0.03, 0.76) | **0.027** | 0.20<br>(0.04, 0.77) | **0.029** | 0.19<br>(0.04, 0.76) | **0.025** |
| Age | 1.07<br>(1.01, 1.13) | **0.028** | 1.06<br>(0.997, 1.12) | 0.071 | 1.07<br>(1.01, 1.14) | **0.018** |
| HHS | 7.89<br>(2.10, 37.3) | **0.004** | 6.79<br>(1.81, 31.8) | **0.008** | 8.22<br>(2.16, 32.8) | **0.004** |
| Sex | 0.81<br>(0.23, 2.90) | 0.75 | 0.52<br>(0.14, 1.81) | 0.32 | 0.70<br>(0.20, 2.43) | 0.58 |
| Race | 0.92<br>(0.28, 3.37) | 0.90 | 1.63<br>(0.47, 6.51) | 0.46 | 1.12<br>(0.34, 4.05) | 0.85 |

Abbreviations: OR (odds ratio), CI (confidence interval), HHS (Hunt Hess Scale score)

### Machine learning (ML) models

ML models were developed to test the ability of clinical variables with or without the addition of metabolites to predict DCI. Prediction models based on ELR using baseline clinical variables showed AUROC ranging from 0.34 to 0.69 with an average AUROC of 0.51 (95% CI: 0.38-0.64) (Figure 5A). Models based on XGB showed similar performance with AUROC ranging from 0.34 to 0.81 with an average AUROC of 0.58 (95% CI: 0.43-0.73) (Figure 5B). Models were also developed including both baseline clinical variables and five metabolites associated with DCI (S-adenosyl homocysteine, undecanedioate, octanoylcarnitine, sphinganine, and sphingosine). Models based on ELR showed AUROC ranging from 0.34 to 0.81 with an average AUROC of 0.79 (95% CI: 0.70-0.88) (Figure 5C), and models based on XGB showed AUROC ranging from 0.69 to 0.97 with an average AUROC of 0.84 (95% CI: 0.72-0.96) (Figure 5D). Including metabolites resulted in significant improvements in AUROC: ELR, *p* < 0.01 (Figure 5A vs5C) and XGB, *p* = 0.016 (Figure 5B vs 5D).

**Figure 5:**
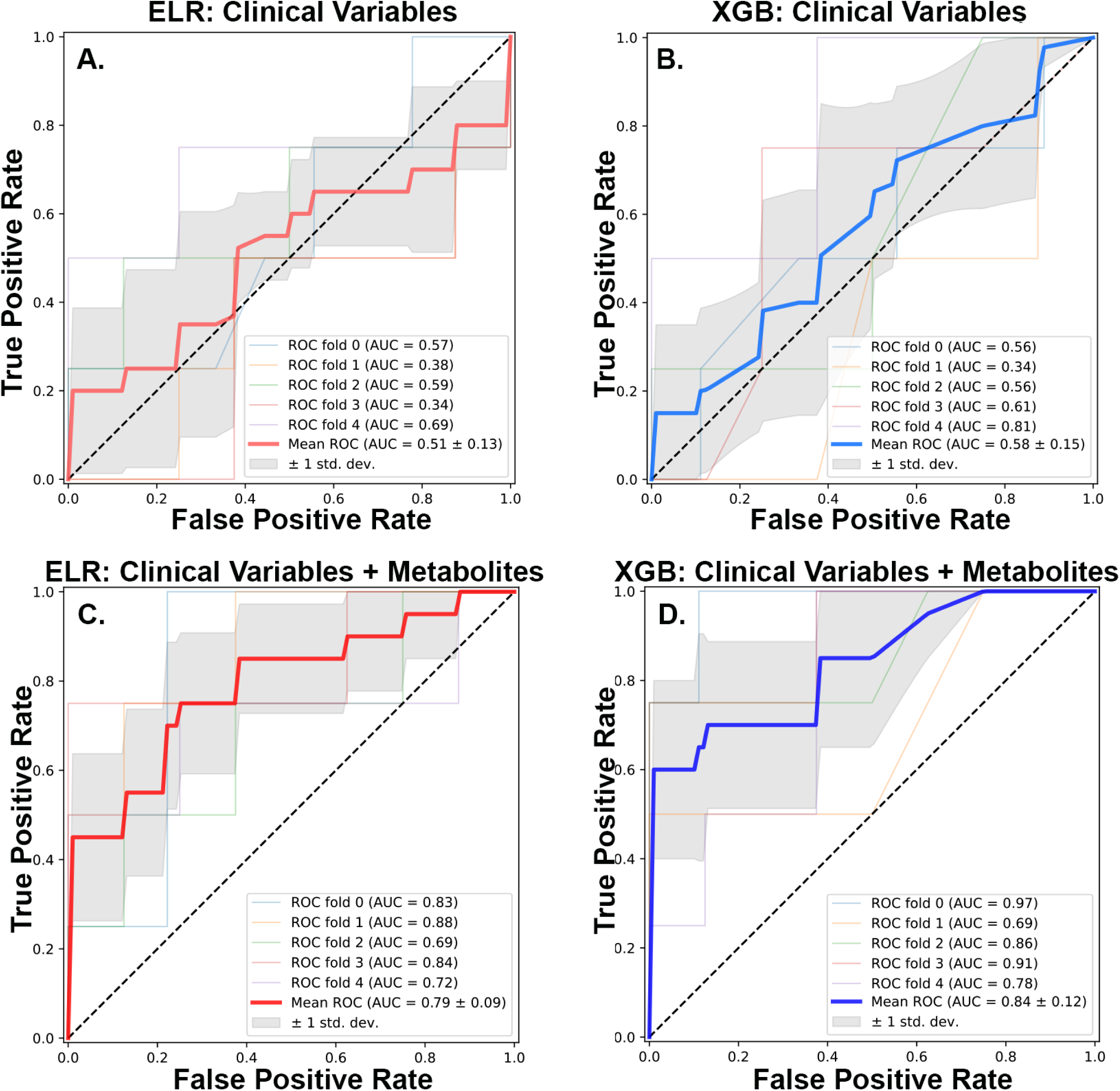
Performance of machine learning models with and without metabolites for prediction of DCI. ELR and XBG machine learning models were developed to predict DCI using clinical variables alone or a combination of clinical variables and metabolites. Compared to the ELR prediction models solely based on clinical variables (A), addition of metabolites to the predictors significantly improves the model’s performance (C) (p < 0.01). A similar improvement was observed when comparing XGB models based on clinical variables (B) vs. clinical variables and the metabolites (D) (p < 0.05). Abbreviations: DCI (Delayed Cerebral Ischemia), ROC (Receiver Operating Characteristic) curve, AUC (Area Under the Curve), ELR (elastic net Linear Regression), XGB (Extreme Gradient Boosting)

For the prediction of DCI, metabolites consistently showed higher importance than age and HHS score. When MDI for different features were compared, all metabolites showed higher feature importance than age and HHS. S-adenosylhomocysteine and undecanedioate showed particularly high importance with MDI of 0.35 and 0.19, respectively, suggesting more than half of the important decision leaf nodes in predicting DCI consisted of these two metabolites (Figure 6). Comparison based on SHAP values also supported the importance of metabolites in predicting DCI. The mean magnitudes of SHAP values for undecanedioate, S-adenosylhomocysteine and sphinganine were higher than the clinical variables, and their influence was consistent throughout their respective ranges of values (Figure 6).

**Figure 6:**
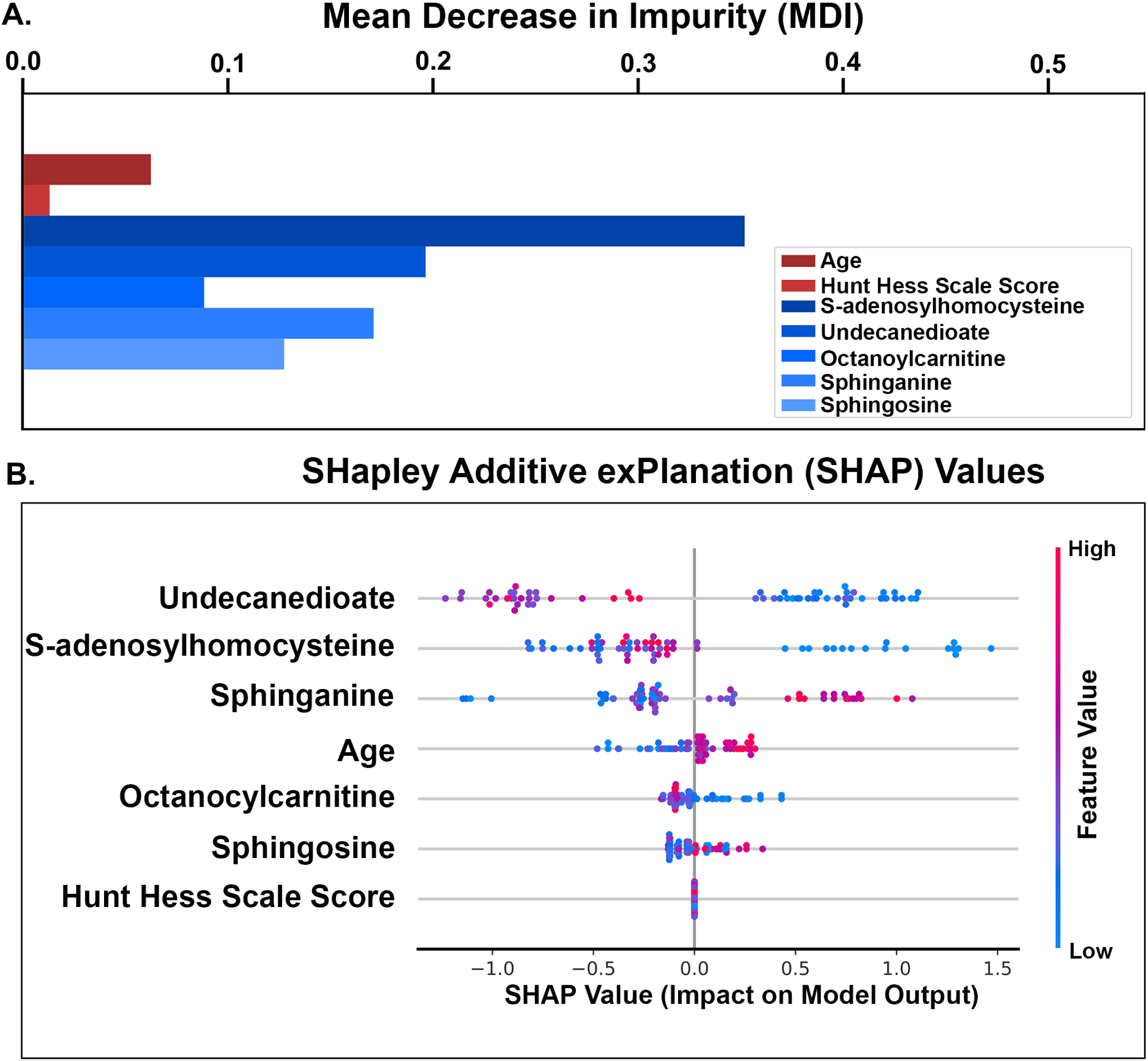
Feature importance in the XGB prediction model for DCI. Feature importances of clinical variables and metabolites in predicting DCI were compared using a mean decrease in impurity (A) and Shapley additive explanation (SHAP) values (B). Both analyses suggest that the metabolites are generally more important than clinical variables in predicting DCI. Abbreviations: DCI (delayed cerebral ischemia), XGB (Extreme Gradient Boosting), HH (Hun Hess) grade,

## Discussion

This study took an untargeted metabolomic approach to comprehensively identify circulating metabolites that are specific to aSAH and able to predict outcomes. We included subjects from two tertiary medical centers and analyzed samples collected both early after aneurysm rupture and after 7 days. Lipid and amino acid metabolites were critical in distinguishing aSAH from control subjects. Circulating FFAs were markedly increased after aSAH, even when compared with critically ill control subjects without neurological injury. Early changes in several metabolites also showed associations with clinical outcomes. Although DCI is notoriously difficult to predict, including metabolites in ML models significantly improved predictive ability.

Although studies have assessed metabolites occurring in the CSF compartment after aSAH^24^, relatively less attention has been given to the circulating metabolome. CSF metabolites may be more reflective of processes occurring in the central nervous system, but they do not provide insight into metabolic factors contributing to the robust systemic inflammatory response occurring after aSAH^25^. We previously demonstrated that TCA cycle metabolites were associated with functional outcomes as well as levels of inflammatory cytokines^8^. These changes occurred before peak cytokine levels^26^, suggesting that circulating metabolites may precede and promote the systemic inflammatory response. A recent study employing lipidomics from plasma samples after acute brain injury (ABI) included 30 aSAH subjects and demonstrated that several lipid metabolites were predictive of outcomes^27^. Plasma metabolites may therefore be excellent biomarkers of outcomes after aSAH and provide insight into the pathophysiological changes contributing to complications such as DCI after aneurysm rupture.

We demonstrated marked shifts in lipid metabolites after aSAH with dramatic increases in FFAs. Alterations in lipid metabolism are not unique to aSAH and occur in other causes of critical illness. For instance, in sepsis, catecholamines activate enzymes such as hormone sensitive lipase (HSL) and adipose triglyceride lipase (ATGL) resulting in increased lipolysis and release of FFAs into the circulation^28^. Although increased CSF lipids have been demonstrated after aSAH^29^, our study is the first to demonstrate the extent of circulating lipid changes occurring after aSAH relative to healthy and critically ill controls. Pathway analysis (Figure S1) revealed that altered lipid metabolites are involved in processes such as ferroptosis, PI3K-Akt signaling, and steroid hormone biosynthesis – all of which have been implicated in aSAH pathophysiology^30,31^. Recently, elevated FFAs have been associated with poor outcomes after ABI^27^.

In addition to changes in lipid metabolites, our results reveal decreases in circulating amino acids in aSAH compared with control subjects. This is consistent with previous results demonstrating a catabolic state occurring after aSAH due to increased catecholamine release and cytokine production^32^. This study included a subset of patients from the UMSOM who were enrolled in the INSPIRE clinical trial, which tested the ability of a high protein diet (HPRO) and neuromuscular electrical stimulation (NMES) to preserve muscle mass after aSAH^13^. All samples from the early time point included in the present study were prior to randomization before any subject had received the intervention. We previously demonstrated that the HPRO+NMES intervention resulted in an increase in amino acid metabolites, with several having significant associations with preserved quadricep and temporalis muscle mass^33^. While these findings are likely not specific to aSAH, aSAH subjects exhibited elevated amino acid metabolism relative to critically ill controls.

Among the metabolites associated with functional outcomes and DCI, acylcarnitines (AC) were particularly notable. Increased ACs were found among both patients with good outcomes and those without DCI. Importantly, these findings were replicated in subjects from two separate institutions – UTHSC and UMSOM (Figure S2, S3). ACs serve as the primary form in which fatty acids are transported into mitochondria for fatty acid β-oxidation (FAO)^34^. In ischemic stroke, increased circulating ACs at presentation were found to be associated with stroke recurrence^35^. Similarly, in patients with TBI, circulating ACs were correlated with increased mortality and poor functional outcomes ^36^. However, production of circulating ACs may also reflect metabolic flexibility and a swift toward FAO. Indeed, temporary increases during fasting or exercise may reflect beneficial metabolic processes^37^. A recent animal study showed that supplementation with carnitine reduced vasospasm after aSAH^38^. Our results demonstrate larger decreases in ACs from T1 to T2 among patients with good outcomes, suggesting increased fatty acid utilization for energy generation via FAO.

Our results reveal a relationship between increased lysophospholipids (LPLs) and improved functional outcomes after aSAH. Similar results have been shown after traumatic brain injury (TBI) of various severity^39,40^, however, this is the first report to link LPLs to functional outcomes after aSAH. Primarily synthesized as breakdown products of plasma membrane phospholipid, LPLs have been hypothesized to be critical in restoring cellular membranes after neuronal injury through conversion into phospholipids necessary for membrane repair by lysophospholipid acyltransferase^41^. However, LPLs can also serve an opposing role as a substrate for proinflammatory LPA production by autotaxin (ATX)^42^. LPA may contribute to both blood brain barrier dysfunction and homing of activated inflammatory cells^43^. It is therefore conceivable that increased LPLs among patients with better outcomes may reflect the diversion of LPLs into reparative pathways rather than LPA production by ATX.

In comparison with functional outcomes, increased sphingolipids (SL) (sphingosine and sphinganine) were found to be associated with the occurrence of DCI. Sphingosine kinase (SK) phosphorylates sphingosine to produce the more bioactive molecule S1P. In aSAH, S1P has been hypothesized to have pathophysiological effects within the brain parenchyma and CSF, with the general belief that CSF S1P contributes to vasospasm^44^. However, the circulating lipid metabolites identified herein may contribute to DCI through their interaction with peripheral inflammatory cells. Circulating monocytes have been shown to traffic to the CNS and contribute to DCI ^45^, and lipid metabolites are pivotal for monocyte activation and migration. LPA is involved in monocyte transformation into macrophages^46^ and acts as a signal for monocyte migration^47^. LPA upregulates monocyte expression of programmed cell death ligand 1 (PD-L1), a key signaling molecule for monocyte trafficking to the CNS after aSAH^47^.

Our results are consistent with previous reports showing that standard models have poor predictive ability for DCI ^48^. Incorporating clinical variables into ML models has been shown to improve the prediction of DCI and vasospasm^48,49^. We demonstrated that incorporating metabolites detected in plasma samples collected early after aSAH into ML models can significantly improve the prediction of DCI. However, these results require verification in larger, independent cohorts of aSAH patients. Although we included subjects from two tertiary care centers (UTHSC and UMSOM), we were unable to develop training and validation cohorts from separate sites due to insufficient sample sizes. Compared with a recent study using lipid metabolites in ML models for the prediction of functional outcomes^27^, our models were specific for the detection of DCI rather than functional outcomes after aSAH and incorporated unique metabolites. Functional outcomes, especially longer term, and heavily influenced by clinical variables such as age and aSAH clinical severity, and we believe that metabolites may play a more important role in the prediction of DCI. Indeed, feature importance for DCI prediction showed that metabolites had an outsized impact on outcomes prediction, in particular S-adenosylhomocysteine. This may suggest imbalances in methylation or redox status^50^.

Our study has important limitations. Firstly, the large number of metabolites detected resulted in a risk of false discovery. Although we employed FDR-corrected p-values for comparisons between aSAH and control subjects, FDR-corrected p-values for metabolites identifying DCI and functional outcomes did not reach the prespecified threshold for statistical significance. Instead, we ranked metabolites based on their uncorrected p-values. These metabolites are therefore especially susceptible to type I error. Second, our analysis did not extensively consider comorbid conditions and medication histories. Metabolites associated with cellular energy metabolism are affected by metabolic diseases and medications that alter metabolism. We were also unable to account for medications and nutritional status during hospitalization. However, no patients were yet taking an oral diet at the time of first sample collection after hospital admission. Third, we have not analyzed the associations between metabolomic profiles and longer-term outcomes. We suspect that the initial metabolomic profile after aSAH is more predictive of early complications with factors such as age, comorbidities, and amount of physical rehabilitation received being more relevant to longer-term outcomes. However, whether relationships exist between specific metabolites and longer-term outcomes will require further studies. Lastly, the untargeted metabolic approach that we took in this study only provides a snapshot of the level of each metabolite identified and provides limited mechanistic insight. Future studies incorporating transcriptomic and proteomic experiments will be required to understand upstream and downstream changes affecting the metabolites we identified in this report.

## Conclusions

aSAH triggers a systemic response that exacerbates early brain injury and causes delayed complications, including DCI and poor functional outcomes. Herein, we performed untargeted metabolomics using plasma samples from aSAH patients and demonstrated significant shifts in lipid and amino acid metabolites compared with controls. Lipid metabolites (LPLs, SLs, and ACs) were predictive of DCI and functional outcomes and improved the performance of ML models. While these metabolites require verification, we believe that they provide important insights into pathophysiological changes occurring after aSAH and may serve as clinically useful biomarkers for risk stratification.

## Data Availability

Data will be shared upon request, contingent on compliance with institutional or ethical guidelines.

## Statements and Declarations

The authors confirm that the manuscript complies with all instructions to the authors.

All authors read and approved the final manuscript before submission.

This article has not been submitted or published elsewhere.

## Acknowledgments

None

## Author contributions

Bosco Seong Kyu Yang, Aaron Gudson: conceptualization, methodology, formal analysis, investigation, writing – original draft, writing – review & editing, visualization

Jude Savarraj, Folefac Atem, Philip Lorenzi, Xuefang Ren, Neeraj Badjatia, Huimahn Choi: validation, writing – review & editing

Hua Chen, Sarah Hinds, Glenda Torres, Alice Ryan: data collection, data curation, formal analysis

